# From Incidence to Inequality: Charting the Global Burden of Testicular Cancer in Adolescents and Young Adults for the Global Burden of Disease Study 2021

**DOI:** 10.1101/2025.06.19.25329967

**Authors:** Hongting Xie, Quan Sun, Peng Xue, Xuelei Chu, Feiyu Xie, Shijie Zhu, Lingze Xi, Jiaming Hong, Xingdong Lin, Jun Lyu, Nanbu Wang

## Abstract

**Background:** The incidence of testicular cancer (TC) among adolescents and young adults (AYAs: aged 15-39 years) population presents unique biological and epidemiologic characteristics, and there is currently a wide variation in incidence and survival across regions.

**Methods:** The data on incidence, prevalence, mortality, and disability-adjusted life-years (DALYs) owing to TC were obtained from the Global Burden of Disease 2021. We quantified differences in the burden of TC among AYA at the socio-demographic index (SDI), regional, and national levels. Decomposition analysis was used to identify the main drivers of variation in the burden. Frontier analysis demonstrated the potential for countries to reduce the burden.

**Results:** Globally, there were 28499 new cases, 61596 prevalent cases, 5699 mortality cases, and 376564 DALYs due to TC among AYA in 2021. Between 1990 and 2021, the age-standardized incidence rate (ASIR) and age-standardized prevalence rate (ASPR) increased while the age-standardized mortality rate (ASMR) and age-standardized DALYs rate (ASDR) decreased. Regionally, Southern Latin America exhibited the highest ASRs. Nationally, Monaco ranked highest in ASIR and ASPR, while Mexico had the highest ASMR and ASDR. Population growth and epidemiological changes were major drivers for the burden of DALYs in middle SDI regions, which had the greatest potential for improvement.

**Conclusions:** The global burden of TC among AYA has risen, which is most pronounced in middle SDI regions. Among the AYA population, TC survivors have increased significantly due to the decreased ASMR and ASDR, who badly need more attention for TC treatment.

## 1. Introduction

In the 21st century, cancer remains a leading contributor to significant social, economic, and public health challenges (1). Although adolescents and young adults (AYA) account for only 5% of the global cancer burden (2), the incidence of cancer within this population has been steadily increasing worldwide (3). The social, cultural, and economic burdens faced by AYA cancer patients during treatment contribute to a markedly higher suicide rate in this population compared to others (4), with testicular cancer (TC) showing the highest suicide rate within the AYA cancer (5).

Previous epidemiological studies have elaborated that Europe, North America, Australia, and New Zealand exhibit the highest TC incidence, while Asian and African countries report significantly lower rates (6). Furthermore, countries with transitioning economies have shown a consistent upward trend in TC incidence over the years (7). Nevertheless, the morbidity-to-mortality ratio in the highly developed Nordic countries is 26:1, in stark contrast to the ratio of 2:1 observed in South-East Asia, South-Central Asia, and Africa (6). Owing to a significant genetic predisposition, increased exposure to risk factors (8), and advancements in early detection and screening technologies (9), the incidence of TC has risen and become the most prevalent malignancy among males in the AYA population (10). Over 95% of TC patients become survivors, largely owing to the implementation of radical surgical interventions, platinum-based chemotherapy regimens, and radiation therapy (11). However, despite these improvements in survival rates, TC survivors often face significant long-term challenges, including temporary or permanent infertility, sexual dysfunction, cardiovascular toxicity from platinum-containing therapies, and depression. These disparities underscore the importance of elucidating the global trends in the incidence, prevalence, mortality, and DALYs of TC among AYA. This knowledge is essential for addressing the inequities in healthcare resource allocation for TC management and improving outcomes across diverse regions.

To address these challenges, this study aims to provide a comprehensive assessment of the burden and epidemiology of TC among AYA by analyzing data from the Global Burden of Disease (GBD) Study 2021 (Injury and Risk Factor Collaborators). We characterized the global epidemiology of TC in individuals aged 15-39 years and examined trends over time. Moreover, we employed decomposition analysis to explore potential factors contributing to changes in TC among AYA burden. Lastly, we analyzed potential improvements in TC among AYA burden to identify countries or regions where additional efforts are needed, to offer valuable references for the prevention and management strategies of TC among AYA.

## 2. Methods

### 2.1. Study population and data collection

This study’s data were taken from GBD 2021, which provides an up-to-date and comprehensive analysis of 371 diseases among 204 countries and territories using recent epidemiological data and improved standardized methods (12). All countries and territories were classified into 21 regions and grouped into five categories based on the socio-demographic index (SDI). The SDI is a comprehensive measure of education, economic, and fertility levels, including five levels corresponding to the five SDI quintiles, namely, low (< 0.47), low-middle (0.47–0.62), middle (0.63–0.71), high-middle (0.72–0.81), and high (> 0.81) (13).

We obtained data on the annual incidence, prevalence, mortality, DALYs, and age-standardized rates (ASRs) for TC aged 15-39 years globally from the Global Health Data Exchange (https://vizhub.healthdata.org/gbd-results/). According to prior research, the 15-39 year age group is categorized as AYA, where TC patients face unique physiological and psychological challenges, including treatment side effects, fertility issues, and so on. Therefore, research that specifically targets their unique needs is required.

### 2.2. Ethical Considerations

For GBD studies, a waiver of informed consent was reviewed and approved by the Institutional Review Board of the University of Washington. All the information about ethical standards is available through the official website (http://www.healthdata.org/gbd/2021).

### 2.3 Statistical analysis

Health Metrics and Evaluation’s general methodologies to the GBD 2021 Study and its major improvements over previous cycles have been explained in the previous studies (14). In this study, a 95% uncertainty interval (UI) for each variable was determined using the 2.5th and 97.5th percentiles of the posterior distribution from 1000 draw values. To make comparisons easier, all rates are presented per 100,000 people. All tests were two-sided, and *P* < 0.05 was considered significant. Incidence and prevalence were modelled using Bayesian disease modelling meta-regression tool (Disease Modelling Meta-Regression; version 2.1). Mortality data were obtained from vital registration systems, verbal autopsies, and population-based cancer registry inputs modeled with mortality-to-incidence ratios(15). DALYs, an optimal measurement of disease burden, were calculated as the sum of years of life lost and years lived with disability. We also described and analyzed the rates of disease incidence, prevalence, mortality, and DALYs adjusted for age, and we looked at these rates globally, regionally, and nationally, by SDI.

Joinpoint regression analysis was performed to assess trends in the AYA testicular cancer burden using Joint Command Line Version. This software tracks trends in data over time and then fits the simplest model possible to the data by connecting several different line segments on a logarithmic scale. Average annual percentage changes (AAPCs) were calculated to assess trends, a geometrically weighted average of the different annual percentage changes from the joinpoint trend analysis, for which weights are equal to the length of each period during the specified time interval (16). The 95% confidence interval (CI) was obtained from the linear regression model. For the AAPC value and 95% CI above zero, the corresponding ASR showed an upward trend and vice versa (17).

We applied the decomposition methods developed by Das Gupta to visually demonstrate the role of the three factors driving changes DALYs between 1990 and 2021 (i.e., aging, population, and epidemiology) (18). Moreover, we used non-parametric data envelope analysis and referenced detailed descriptions in previous studies to produce a nonlinear frontier (19). This frontier implies the lowest achievable burden determined according to development status. The detailed methodologies for decomposition and frontier analysis are provided in Supplementary Methods. All the above statistical analyses and graphs were performed using R version 4.3.2.

## 3. Results

### 3.1 Global trends

There was an evident rise in TC incidence and prevalence among the AYA population. For incidence, there were 28,499 (95% UI: 27,000 to 30,124) new cases in 1990 compared to 61,596 (95% UI: 57,749 to 66,174) new cases in 2021, which rose 53.73%. Over the past 32 years, there was a 55.80% rise in prevalence from 209,012 (95% UI: 195,998 to 223,138) cases to 472,966 (95% UI: 440,012 to 512,848). Regarding ASR, the global TC among the AYA population age-standardized incidence rate (ASIR) and age-standardized prevalence rate (ASPR) in 2021 were 4.05 (95% UI: 3.8 to 4.35)/100,000 and 31.05 (95% UI: 28.88 to 33.68)/100,000, with a significant upward trend from 1990 to 2021(ASIR, AAPC = 1.51%; 95% CI: 1.13 to 1.9; ASPR, AAPC = 1.73%; 95% CI: 1.64 to 1.81), respectively.

As shown in Fig. 1A, the global ASIR showed significant increases after a slow decline in 1990-1993 (APC = −0.46% [95 % CI, −2.47 to 1.59], *P >* 0.05), with marked increases in 1993-1997 (APC = 3.19% [95% CI, 1.07 to 5.35], *P* < 0.05) and 2002-2021 (APC = 1.80% [95% CI, 1.67 to 1.93], *P* < 0.05). Additionally, the global ASPR has markedly increased between 1990 and 2021 (APC = 1.73% [95% CI, 1.64 to 1.81], *P* < 0.05) (Fig. 1B).

**Fig. 1.**
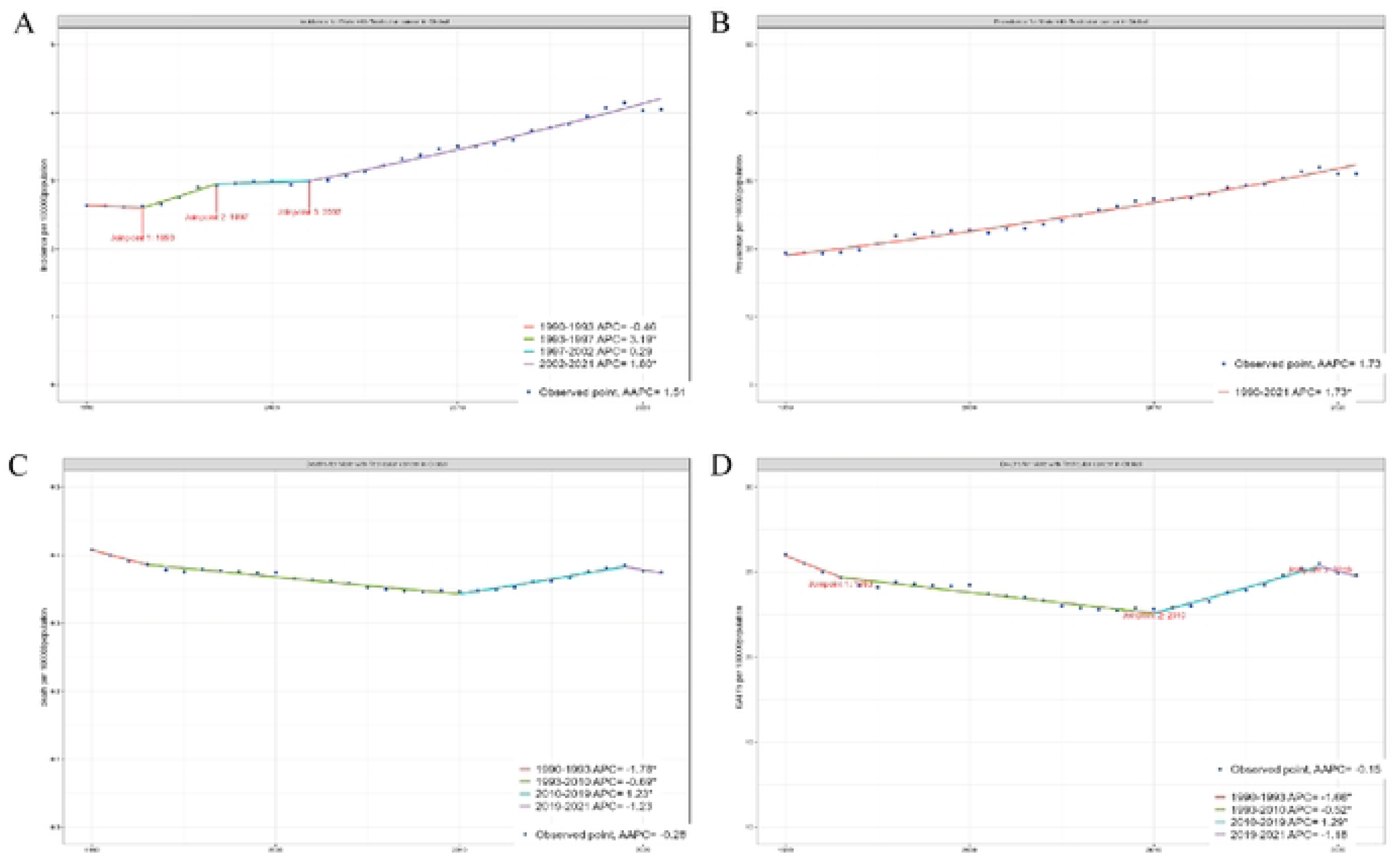
Global trends in age-standardized rates of testicular cancer among adolescents and young adults from 1990 to 2021. (A) Age-standardized incidence rate. (B) Age-standardized prevalence rate. (C) Age-standardized mortality rate. (D) Age-standardized DALYs rate. Abbreviations: AAPC average annual percent change; APC annual percent change; DALYs disability-adjusted life years.

From 1990 to 2021, the number of worldwide mortality associated with TC among the AYA population increased from 4,400 (95% UI: 4,099 to 4,736) to 5,699 (95% UI: 5,288 to 6,156) with a 22.79% rise (Table 1). Moreover, the number of DALYs increased from 282,637 (95% UI: 262,651 to 304,571) to 376,564 (95% UI: 348,929 to 408,401) (Table 1).

**Table 1.**
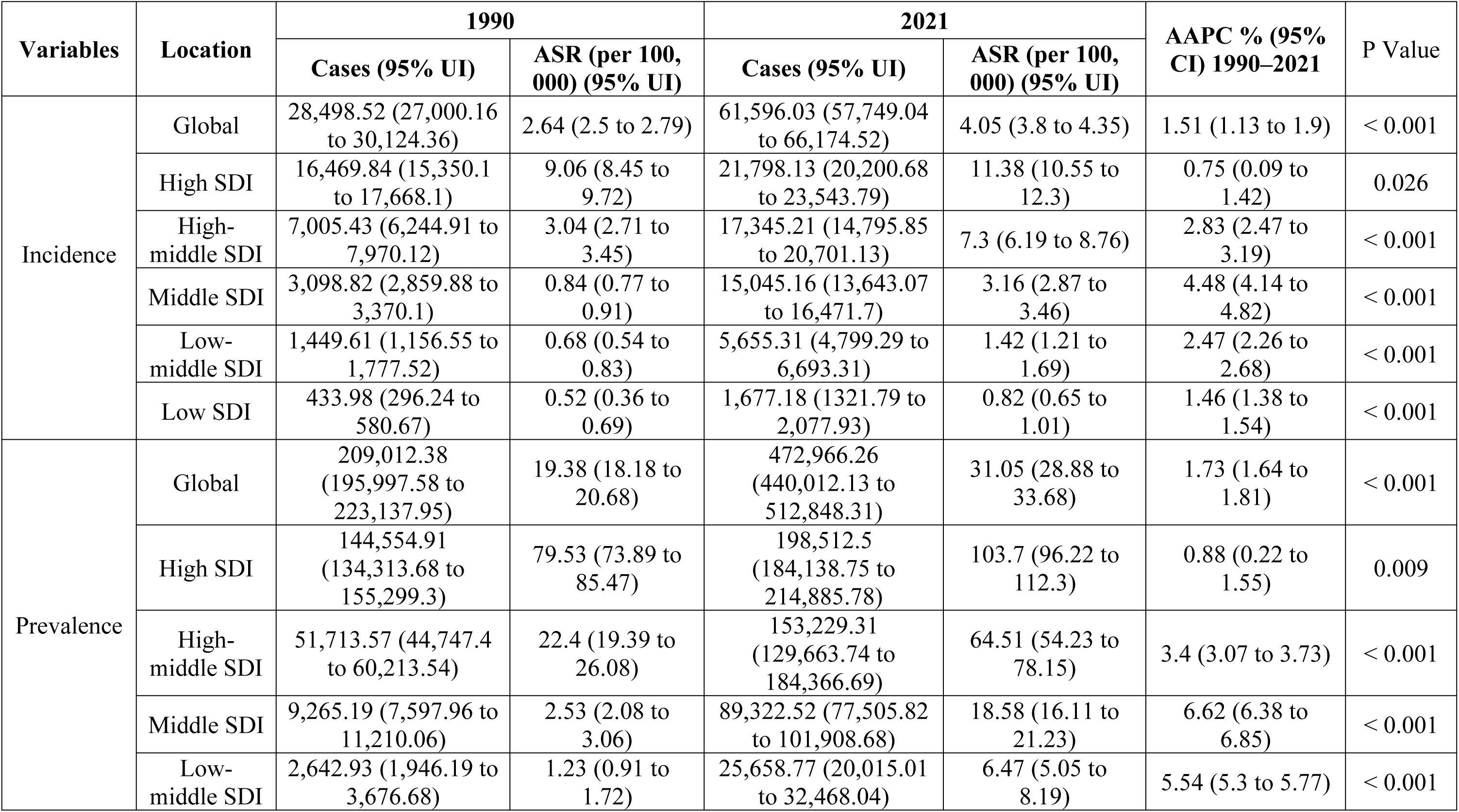
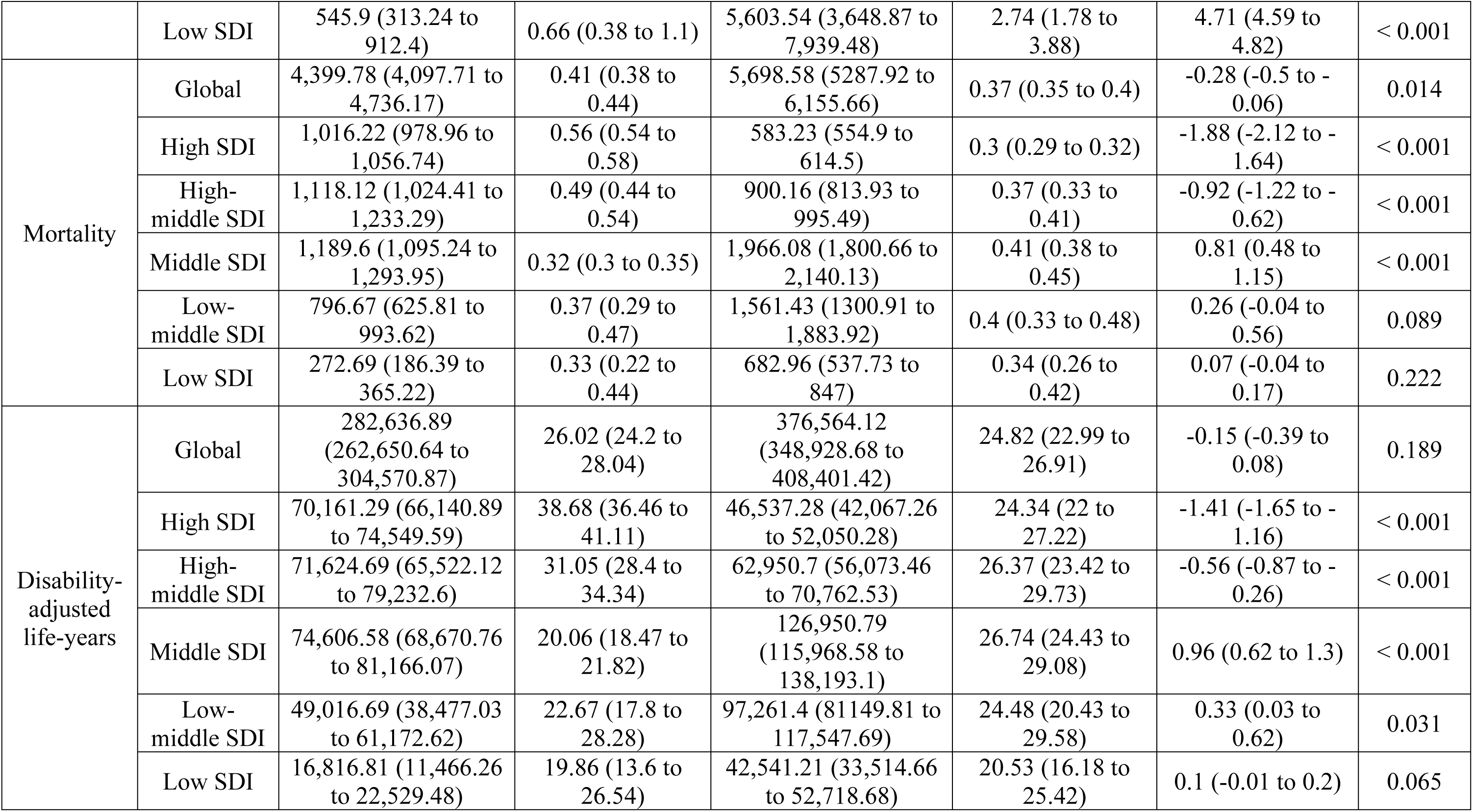
The global incidence, prevalence, mortality, and disability-adjusted life-years of testicular cancer in adolescents and young adults, with AAPC from 1990 and 2021.

| Variables | Location | 1990 |  | 2021 |  | AAPC % (95% CI) 1990–2021 | P Value |
| --- | --- | --- | --- | --- | --- | --- | --- |
|  |  | Cases (95% UI) | ASR (per 100,000) (95% UI) | Cases (95% UI) | ASR (per 100,000) (95% UI) |  |  |
| Incidence | Global | 28,498.52 (27,000.16 to 30,124.36) | 2.64 (2.5 to 2.79) | 61,596.03 (57,749.04 to 66,174.52) | 4.05 (3.8 to 4.35) | 1.51 (1.13 to 1.9) | < 0.001 |
|  | High SDI | 16,469.84 (15,350.1 to 17,668.1) | 9.06 (8.45 to 9.72) | 21,798.13 (20,200.68 to 23,543.79) | 11.38 (10.55 to 12.3) | 0.75 (0.09 to 1.42) | 0.026 |
|  | High-middle SDI | 7,005.43 (6,244.91 to 7,970.12) | 3.04 (2.71 to 3.45) | 17,345.21 (14,795.85 to 20,701.13) | 7.3 (6.19 to 8.76) | 2.83 (2.47 to 3.19) | < 0.001 |
|  | Middle SDI | 3,098.82 (2,859.88 to 3,370.1) | 0.84 (0.77 to 0.91) | 15,045.16 (13,643.07 to 16,471.7) | 3.16 (2.87 to 3.46) | 4.48 (4.14 to 4.82) | < 0.001 |
|  | Low-middle SDI | 1,449.61 (1,156.55 to 1,777.52) | 0.68 (0.54 to 0.83) | 5,655.31 (4,799.29 to 6,693.31) | 1.42 (1.21 to 1.69) | 2.47 (2.26 to 2.68) | < 0.001 |
|  | Low SDI | 433.98 (296.24 to 580.67) | 0.52 (0.36 to 0.69) | 1,677.18 (1321.79 to 2,077.93) | 0.82 (0.65 to 1.01) | 1.46 (1.38 to 1.54) | < 0.001 |
| Prevalence | Global | 209,012.38 (195,997.58 to 223,137.95) | 19.38 (18.18 to 20.68) | 472,966.26 (440,012.13 to 512,848.31) | 31.05 (28.88 to 33.68) | 1.73 (1.64 to 1.81) | < 0.001 |
|  | High SDI | 144,554.91 (134,313.68 to 155,299.3) | 79.53 (73.89 to 85.47) | 198,512.5 (184,138.75 to 214,885.78) | 103.7 (96.22 to 112.3) | 0.88 (0.22 to 1.55) | 0.009 |
|  | High-middle SDI | 51,713.57 (44,747.4 to 60,213.54) | 22.4 (19.39 to 26.08) | 153,229.31 (129,663.74 to 184,366.69) | 64.51 (54.23 to 78.15) | 3.4 (3.07 to 3.73) | < 0.001 |
|  | Middle SDI | 9,265.19 (7,597.96 to 11,210.06) | 2.53 (2.08 to 3.06) | 89,322.52 (77,505.82 to 101,908.68) | 18.58 (16.11 to 21.23) | 6.62 (6.38 to 6.85) | < 0.001 |
|  | Low-middle SDI | 2,642.93 (1,946.19 to 3,676.68) | 1.23 (0.91 to 1.72) | 25,658.77 (20,015.01 to 32,468.04) | 6.47 (5.05 to 8.19) | 5.54 (5.3 to 5.77) | < 0.001 |
|  | Low SDI | 545.9 (313.24 to 912.4) | 0.66 (0.38 to 1.1) | 5,603.54 (3,648.87 to 7,939.48) | 2.74 (1.78 to 3.88) | 4.71 (4.59 to 4.82) | < 0.001 |
| Mortality | Global | 4,399.78 (4,097.71 to 4,736.17) | 0.41 (0.38 to 0.44) | 5,698.58 (5287.92 to 6,155.66) | 0.37 (0.35 to 0.4) | -0.28 (-0.5 to -0.06) | 0.014 |
|  | High SDI | 1,016.22 (978.96 to 1,056.74) | 0.56 (0.54 to 0.58) | 583.23 (554.9 to 614.5) | 0.3 (0.29 to 0.32) | -1.88 (-2.12 to -1.64) | < 0.001 |
|  | High-middle SDI | 1,118.12 (1,024.41 to 1,233.29) | 0.49 (0.44 to 0.54) | 900.16 (813.93 to 995.49) | 0.37 (0.33 to 0.41) | -0.92 (-1.22 to -0.62) | < 0.001 |
|  | Middle SDI | 1,189.6 (1,095.24 to 1,293.95) | 0.32 (0.3 to 0.35) | 1,966.08 (1,800.66 to 2,140.13) | 0.41 (0.38 to 0.45) | 0.81 (0.48 to 1.15) | < 0.001 |
|  | Low-middle SDI | 796.67 (625.81 to 993.62) | 0.37 (0.29 to 0.47) | 1,561.43 (1300.91 to 1,883.92) | 0.4 (0.33 to 0.48) | 0.26 (-0.04 to 0.56) | 0.089 |
|  | Low SDI | 272.69 (186.39 to 365.22) | 0.33 (0.22 to 0.44) | 682.96 (537.73 to 847) | 0.34 (0.26 to 0.42) | 0.07 (-0.04 to 0.17) | 0.222 |
| Disability-adjusted life-years | Global | 282,636.89 (262,650.64 to 304,570.87) | 26.02 (24.2 to 28.04) | 376,564.12 (348,928.68 to 408,401.42) | 24.82 (22.99 to 26.91) | -0.15 (-0.39 to 0.08) | 0.189 |
|  | High SDI | 70,161.29 (66,140.89 to 74,549.59) | 38.68 (36.46 to 41.11) | 46,537.28 (42,067.26 to 52,050.28) | 24.34 (22 to 27.22) | -1.41 (-1.65 to -1.16) | < 0.001 |
|  | High-middle SDI | 71,624.69 (65,522.12 to 79,232.6) | 31.05 (28.4 to 34.34) | 62,950.7 (56,073.46 to 70,762.53) | 26.37 (23.42 to 29.73) | -0.56 (-0.87 to -0.26) | < 0.001 |
|  | Middle SDI | 74,606.58 (68,670.76 to 81,166.07) | 20.06 (18.47 to 21.82) | 126,950.79 (115,968.58 to 138,193.1) | 26.74 (24.43 to 29.08) | 0.96 (0.62 to 1.3) | < 0.001 |
|  | Low-middle SDI | 49,016.69 (38,477.03 to 61,172.62) | 22.67 (17.8 to 28.28) | 97,261.4 (81149.81 to 117,547.69) | 24.48 (20.43 to 29.58) | 0.33 (0.03 to 0.62) | 0.031 |
|  | Low SDI | 16,816.81 (11,466.26 to 22,529.48) | 19.86 (13.6 to 26.54) | 42,541.21 (33,514.66 to 52,718.68) | 20.53 (16.18 to 25.42) | 0.1 (-0.01 to 0.2) | 0.065 |

Regarding ASR, the age-standardized mortality rate (ASMR) and the age-standardized DALYs rate (ASDR) in 2021 were 0.37 (95% UI: 0.35 to 0.4)/100,000 and 24.82 (95% UI: 22.99 to 26.91)/100,000, with slightly decreasing trend over the same period (ASMR, AAPC = −0.28%; 95% CI: −0.50 to −0.06; ASDR, AAPC = −0.15%; 95% CI: −0.39 to 0.08), respectively. More specifically, as illustrated in Fig. 1C and 1D, the global ASMR and ASDR have significantly decreased in 1993-2010 and 2019-2021, with the most notable decreases were observed between 1990 and 1993 (ASMR: APC = −1.78% [95 % CI, −3.08 to −0.46], *P* < 0.05; ASDR: APC = −1.66% [95% CI, −2.97 to −0.32], *P* < 0.05) while slowly increasing in 2010–2019 (ASMR: APC = 1.23% [95% CI, 0.98 to 1.48]; ASDR: APC = 1.29% [95% CI, 1.02 to 1.55], *P* < 0.05).

### 3.2 Global trends by SDI

In 2021, the high SDI region had the most incidence (21,798; 95% UI: 20,201 to 23,544) and prevalence (198,513; 95% UI: 184,139 to 214,886) cases (Table 1), as well as ASRs (ASIR: 11.38; 95% UI: 10.55 to 12.30/100,000, ASDR: 103.7; 95% UI: 96.22 to 112.3/100,000) (Fig. 2A-2B). In addition, the middle SDI region had the most mortality cases (1,966; 95% UI: 1,801 to 2,140) and the most DALYs cases (126,951; 95% UI: 115,969 to 138,193) (Table 1). Likewise, the middle SDI regions had the highest age-standardized mortality (0.41; 95% UI: 0.38 to 0.45/100,000) and DALYs (26.74; 95% UI: 24.43 to 29.08/100,000) (Fig. 2C-2D).

**Fig. 2.**
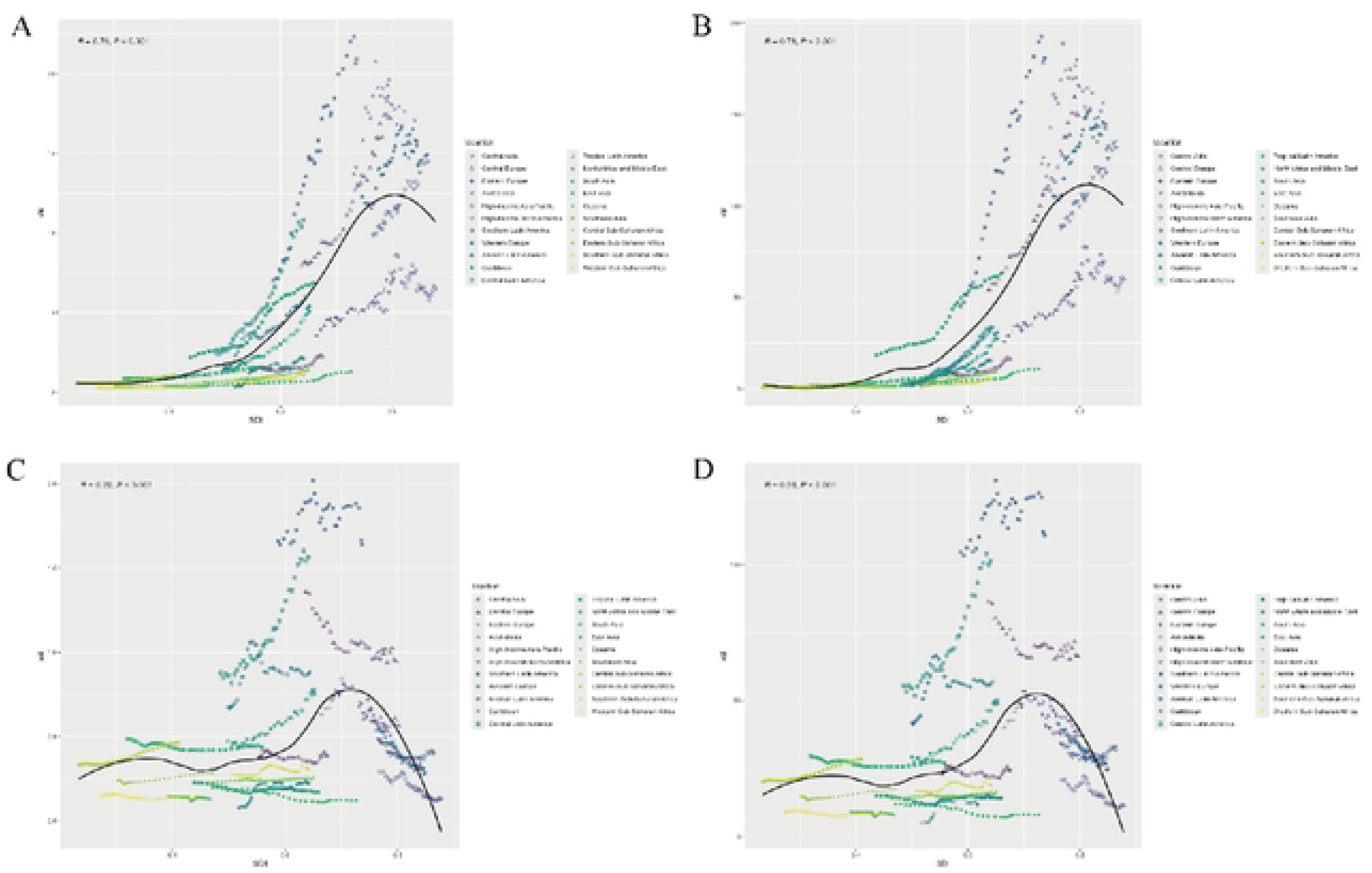
Trends for age-standardized rates of testicular cancer among adolescents and young adults among 21 regions by SDI combined from 1990 to 2021. (A) Age-standardized incidence rate. (B) Age-standardized prevalence rate. (C) Age-standardized mortality rate. (D) Age-standardized DALYs rate. Abbreviations: DALYs disability-adjusted life-years, SDI socio-demographic index.

Interestingly, over the last 32 years, middle SDI regions have shown the fastest growth in ASIR (AAPC = 4.48%; 95% CI: 4.14 to 4.82), ASPR (AAPC = 6.62%; 95% CI: 6.38 to 6.85), ASMR (AAPC = 0.81%; 95% CI: 0.48 to 1.15), and ASDR (AAPC = 0.96%; 95% CI: 0.62 to 1.3), while high SDI regions have shown the slowest or even negative growth in ASIR (AAPC = 0.75%; 95% CI: 0.09 to 1.42), ASPR (AAPC = 0.88%; 95% CI: 0.22 to 1.55), ASMR (AAPC = −1.88%; 95% CI: −2.12 to −1.64), and ASDR (AAPC = −1.41%; 95% CI: −1.65 to −1.16). (Table 1).

### 3.3 Regional trends

In 2021, the region with the highest number of incidence (9,933; 95% UI: 8,623 to 11,509) and prevalence (91,302; 95% UI: 79,230 to 105,941) of TC among the AYA population was Western Europe (Table S1-S2). Besides, South Asia had the most mortality cases (1740; 95% UI: 1429 to 2115) and the most DALYs cases (107817; 95% UI: 88503 to 131204). Surprisingly, Southern Latin America had the highest ASIR (20.93; 95% UI: 15.86 to 27.33/100,000), ASPR (181.03; 95% UI: 130.83 to 238.04/100,000) ASMR (1.64; 95% UI: 1.33 to 1.99/100,000), and ASDR (110.66; 95% UI: 88.83 to 134.71/100,000) (Table S3-S4). During the 32 years, Caribbean had the fastest growth rate in ASIR (AAPC = 6.12%; 95% CI: 4.45 to 7.82), ASMR (AAPC = 3.55; 95% CI: 2.22 to 4.9), and ASDR (AAPC = 3.67; 95% CI: 2.33 to 5.03), but Central Latin America had the fastest growth rate in ASPR (AAPC = 8.88%; 95% CI: 8.31 to 9.45). Respectively, Australasia, High-income Asia Pacific, Western Europe, and Oceania had lower ASIR, ASPR, ASMR, and ASDR (Table S1-S4).

### 3.4 National trends

Among 204 countries in 2021, in AYA population, United States of America had the most TC incidence (7,856; 95% UI: 7,232 to 8,538) and prevalence (71,475; 95% UI: 65,665 to 77,662), while Monaco had the highest age-standardized incidence (86.36; 95% UI: 39 to 167.51/100,000) and prevalence (396.65; 95% UI: 179.35 to 768.37/100,000). In addition, India had the most mortality cases (1,091; 95% UI: 902 to 1,309) and DALYs cases (67,621; 95% UI: 55,897 to 81,032), while Mexico had the highest ASMR (2.31; 95% UI: 2.06 to 2.57) and ASDR (149.33; 95% UI: 133.44 to 166.2) (Table S5-S8, Fig. 3).

**Fig. 3.**
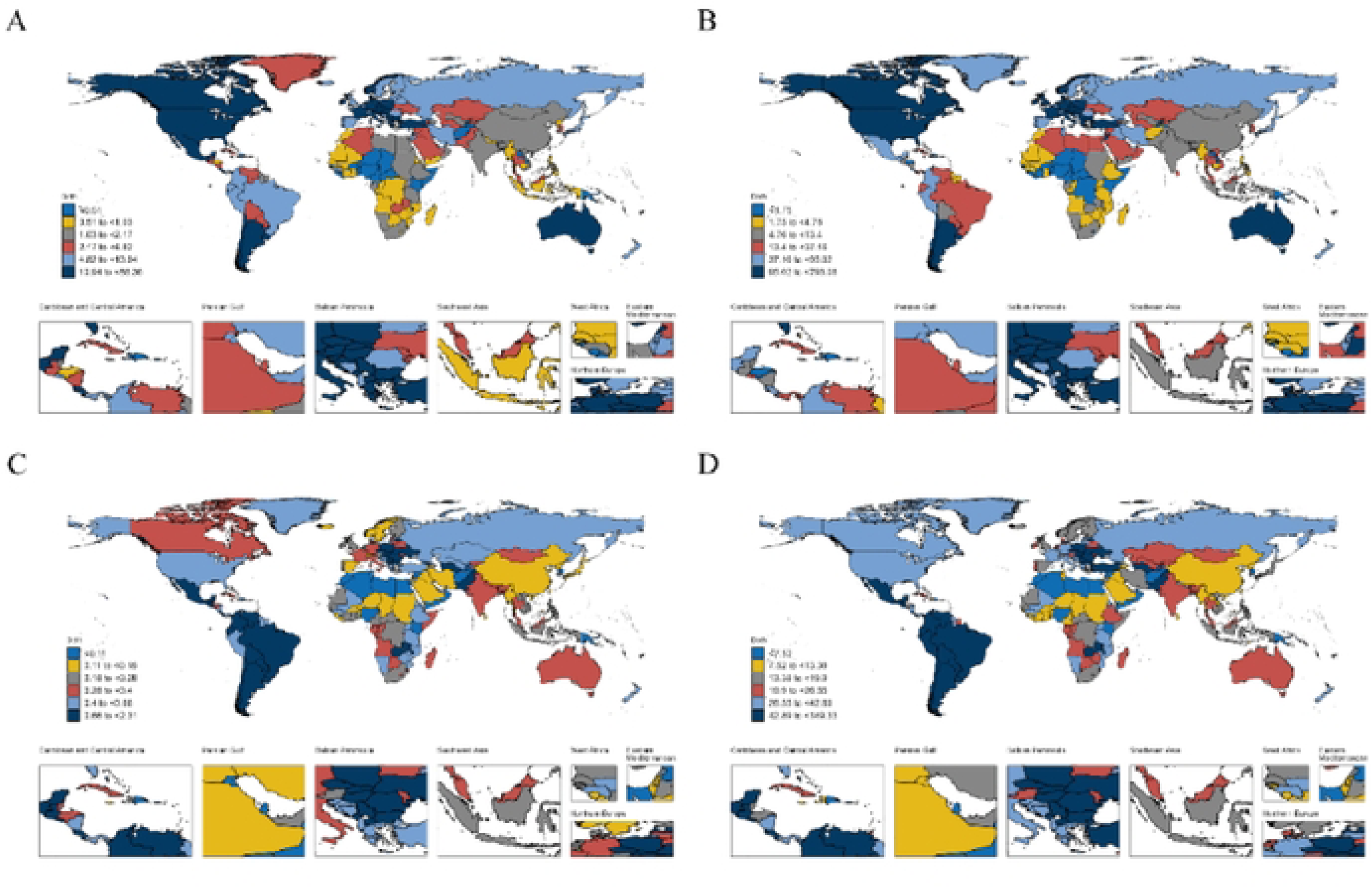
World maps depicting the national age-standardized rates of testicular cancer among adolescents and young adults in 2021. (A) Age-standardized incidence rate. (B) Age-standardized prevalence rate. (C) Age-standardized mortality rate. (D) Age-standardized DALYs rate.

Over the past 32 years, more than 105 countries have shown increasing trends in age-standardized incidence, prevalence, mortality and DALYs rates of TC among AYAs. Puerto Rico showed the highest increase in incidence (AAPC = 9.69%; 95% CI: 7.75 to 11.66), Ecuador had the fastest growth in prevalence (AAPC = 13.64%; 95% CI: 10.65 to 16.72), whereas Belize showed the fastest increase in mortality (AAPC = 7.33%; 95% CI: 4.9 to 9.82) and DALYs (AAPC = 7.17%; 95% CI: 4.94 to 9.44) (Table S5-S8, Fig. 4).

**Fig. 4.**
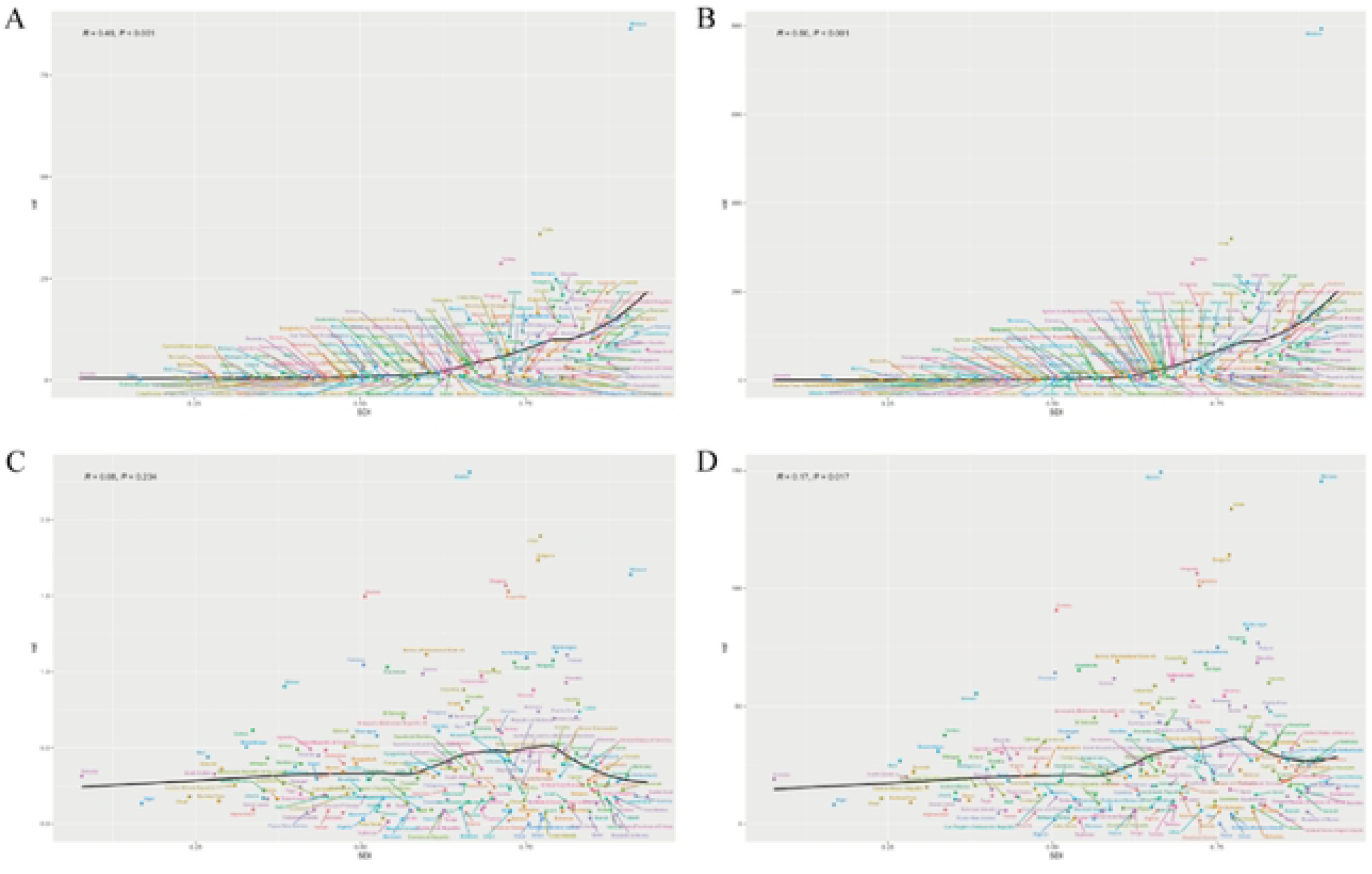
The age-standardised rates of testicular cancer among adolescents and young adults among 204 countries and territories by SDI in 2021. (A) Age-standardized incidence rate. (B) Age-standardized prevalence rate. (C) Age-standardized mortality rate. (D) Age-standardized DALYs rate. Abbreviations: SDI socio-demographic index.

### 3.5 Decomposition analysis of age-standardized DALYs rate

Fig. 5 presents the most significant growth in global DALYs was in the middle SDI quintile regions, while the high and high-middle SDI showed negative growth. Population growth is a driver for the burden of DALYs in Global, Middle, Low-Middle, and low SDI regions. Furthermore, epidemiological change is a growth driver for the burden of DALY in Middle and Middle-low SDI regions, as well as a negative growth factor in Global, High SDI, and High-middle SDI regions (Fig. 5). Globally, aging, population, and epidemiological changes accounted for 8.50%, 107.86%, and −16.37% of the DALY increase (Table S9). The most significant contribution of population growth occurred in the low SDI quintile (95.86%), the highest epidemiological contribution observed in the high-middle SDI quintile (126.06%), and the largest contribution of aging change happened in the middle SDI quintile (5.12%) (Table S9).

**Fig. 5.**
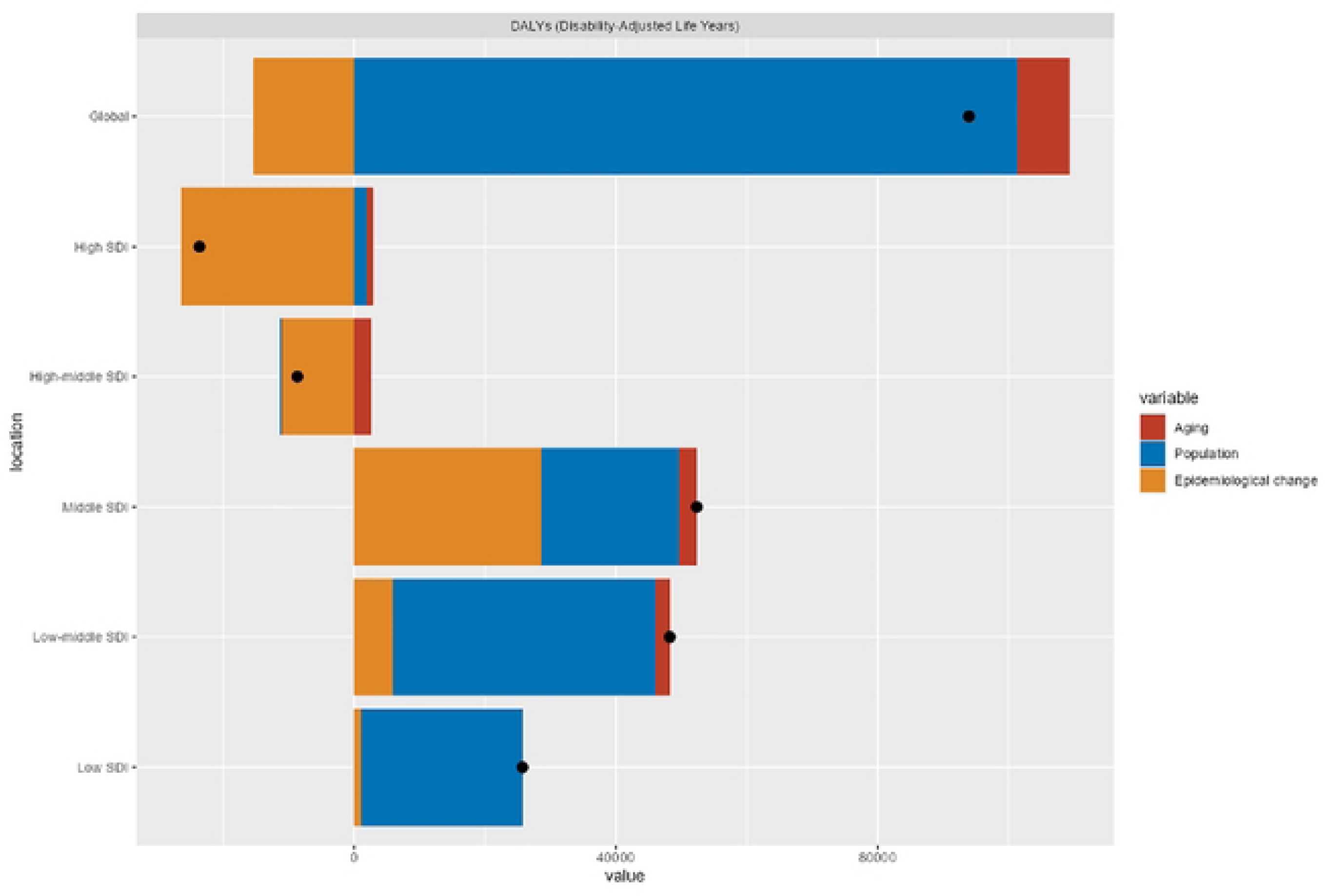
Changes in DALYs of testicular cancer among AYA according to population-level determinants of population growth, aging, and epidemiological change from 1990 to 2021 at the global level and by SDI quintile. Note: The black dot represents the overall value of change contributed by all 3 components. For each component, the magnitude of a positive value indicates a corresponding increase in testicular cancer among AYA attributed to the component; the magnitude of a negative value indicates a corresponding decrease in testicular cancer among AYA attributed to the related component. Abbreviations: AYA: adolescents and young adults, DALYs disability-adjusted life-years, SDI socio-demographic index.

### 3.6 Frontier analysis on the basis of age-standardized DALYs rate

Based on the ASDR and SDI using data from 1990 to 2021, frontier analysis was conducted to illustrate unrealized health benefits for countries or territories at different levels of development (Fig. 6). Fig. 6B and Table S10 provide the effective difference in countries with different SDI in 2021, with an inverted triangle of relationships between them. The countries with middle and high-middle SDI trended to present higher effective differences, while the countries with lower and higher SDI showed lower effective differences. Specifically, the country with the largest effective difference was Mexico (148.83), in the middle SDI quintile, which may have unrealized opportunities for improvement given its resources. Nevertheless, the leading performances were not restricted to developed countries, some countries with low SDI also approached the frontier, such as Nigeria (0). Similarly, Monaco, a high SDI country, with disappointing performance, had a high effective difference (145.47).

**Fig. 6.**
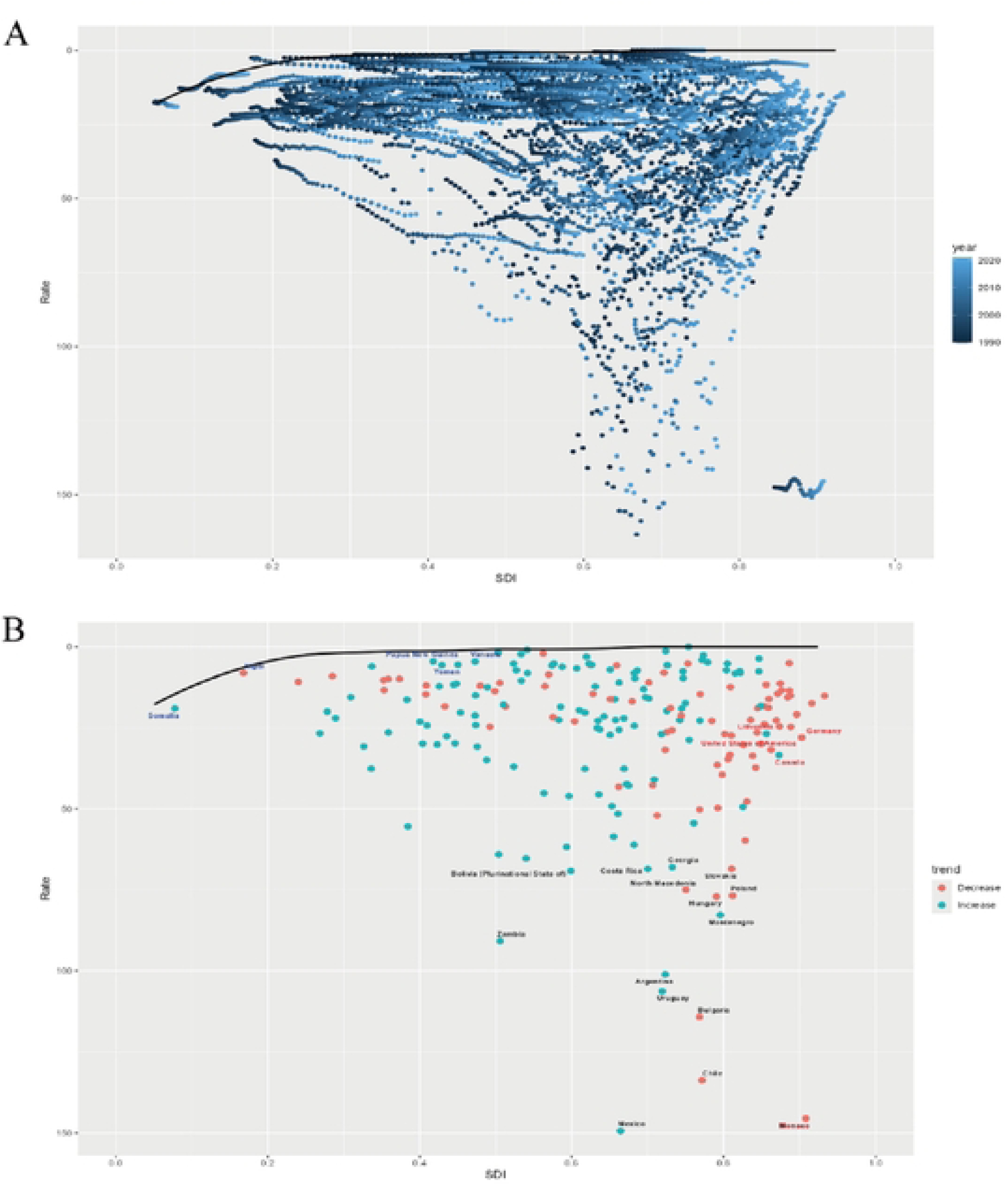
Frontier analysis based on SDI and age-standardized DALYs rate of testicular cancer among AYA from 1990 to 2021. Note: The frontier line was delineated in black. Countries and territories were represented as dots. The color scale ranges from dark blue for 1990 to light blue for 2021. (A). The green dots indicate an increase in the age-standardised DALYs rate for testicular cancer among AYA from 1990 to 2021, while the red dots show a decrease. (B). The top 15 countries with the largest effective difference are highlighted in black. As examples, frontier countries with a lower SDI (< 0.5) and a smaller effective difference are marked in blue, while countries and territories with a higher SDI (> 0.85) and a relatively larger effective difference are marked in red. Abbreviations: AYA: adolescents and young adults, DALYs: disability-adjusted life year, SDI: socio-demographic index.

## 4. Discussion

### 4.1 Global burden of TC in the AYA population

Our study demonstrated a linear increase in the incidence, prevalence, ASIR, and ASPR of TC in the AYA population globally from 1990 to 2021. The rising incidence and prevalence of TC in this population may be closely associated with a variety of factors, including the increasing prevalence of cryptorchidism (20), genetic predisposition, environmental and industrial exposures, and poor lifestyle choices. Among these, cryptorchidism has been identified as the most significant congenital risk factor for TC (21). Similar to cryptorchidism, abdominal plugging in sex reassignment surgeries to reduce genital bulging alters testicular positioning and has also been linked to an elevated risk of TC (22). Additionally, genetic susceptibility plays a crucial role, with specific genes such as CHEK2 (23), KITLG (24), and MTHFR (25) being associated with TC risk. Exposure to endocrine-disrupting chemicals, including elevated levels of androstenedione and Diethylstilbestrol during pregnancy, as well as high androstenedione levels during adolescence, has also been identified as a risk factor for TC (26, 27). Regarding lifestyle factors, similar to other cancers, the consumption of processed meats, alcohol, and tobacco use are recognized as risk factors for TC (8). Moreover, recent studies have provided further evidence that marijuana use is strongly associated with an increased incidence of TC, with the dual use of marijuana and alcohol contributing to even higher cancer risk (28).

In contrast to the sharp increase in the incidence and prevalence of TC, the trend in TC-related deaths within the AYA population has shown a slower rise from 1990 to 2021, with a concomitant decline in the ASMR. This suggests that patients with TC are experiencing improved survival outcomes, which has been reported to have the best cure rates among cancers (29), correlating with an increasing number of AYA survivors. These improvements are largely attributed to advancements in early screening and diagnosis, as well as therapeutic innovations. For instance, more than 80% of patients with stage I TC are cured through radical surgical treatment (30). Moreover, stage II and III TC can also be effectively managed with platinum-based chemotherapy and radiotherapy following surgery, leading to improved overall survival and reduced recurrence rates (31). While aggressive and effective treatments have led to better survival rates, they have also contributed to the growing population of TC survivors. Our study reveals that DALYs in the AYA population have exhibited a gradual upward trend over the past 32 years, indicating that long-term management of TC survivors remains a critical issue in contemporary healthcare (7).

For follow-up medical management, infertility is a significant concern for TC survivors, particularly within the AYA, which exhibits the highest prevalence of TC and represents the optimal reproductive age group (18-39 years). Pre-treatment sperm cryopreservation plays a vital role in preserving fertility for these individuals, regardless of cancer stage or treatment approach (32). While surgical intervention and radiotherapy can adversely affect sperm quality, recovery is often observed within 2-3 years post-treatment (33), indicating that erectile dysfunction (ED) could be a contributing factor to infertility. Approximately 40% of TC survivors report sexual dissatisfaction (34) and experience ED (35). However, the researchers established that on penile Doppler ultrasound, erectile hemodynamics were normal in all patients and 88% responded to phosphodiesterase type 5 inhibitor (36). This reveals that psychological factors, such as anxiety and depression, may play a pivotal role in erectile dysfunction, ultimately affecting sexual health and quality of life. Psychological counseling and comprehensive sexual education are essential components of long-term care for TC survivors. Moreover, platinum-based chemotherapy has been associated with an elevated risk of cardiovascular complications in TC survivors, particularly during the first year following treatment. This includes an increased incidence of vascular thromboembolism, cerebrovascular accidents, myocardial infarctions, and cardiovascular-related mortality (37, 38). Despite the improved survival rates for TC, many young survivors are lost to follow-up for cardiovascular monitoring (38), resulting in insufficient attention. As TC survival rates continue to rise, it is essential to address the multifaceted health challenges faced by survivors, including fertility, sexual health, psychological well-being, and cardiovascular disease. These considerations are crucial for guiding comprehensive follow-up care for the AYA TC population.

### 4.2 Regional and national burden of TC in the AYA population

The burden of TC in the AYA population varies across regions and countries. In general, both the incidence and prevalence of TC increase in higher SDI regions. Western Europe and High-Income North America exhibit the highest incidence and prevalence within the AYA population. These regions are predominantly composed of Caucasian populations, who have a higher prevalence of TC compared to individuals of Asian or African descent (39, 40). A recent study identified 79 single nucleotide polymorphisms associated with TC development, with allele frequencies significantly higher in men of European ancestry compared to those of African descent (41). Cryptorchidism, another major risk factor for TC, is also more prevalent in European and American populations (42). Furthermore, Europe and the United States entered the industrial era earlier and have had prolonged exposure to chemical pollutants. Industrialization, lifestyle habits, and environmental exposures prevalent in these high-SDI regions lead to the elevated incidence and prevalence of TC (8). For example, our study found that the United States has the highest incidence and prevalence of TC among 204 countries. Additionally, another study indicated that TC incidence is rising across all racial and ethnic groups in the U.S. (43). Beyond ethnicity, genetics, and industrial exposure, the legalization of recreational marijuana use in the U.S. might account for the increasing incidence of TC (28). Currently, 24 U.S. states and the District of Columbia have legalized recreational marijuana, with 35.4% of Americans aged 18-25 and 17.2% of those aged 26 or older reporting marijuana use in the past year (28). Repeated use of marijuana, tobacco, and alcohol has been suggested as a potential risk factor for the development of TC in the AYA population.

In 2021, lower SDI regions exhibited higher DALYs and mortality rates in the AYA population. Specifically, South Asia, Central Latin America, and Southeast Asia experienced the highest DALYs and deaths, with India, Mexico, and Pakistan recording the highest number of fatalities at the country level. While patients with early and intermediate-stage TC generally have a favorable prognosis following treatment, the high mortality and DALY rates in the aforementioned regions may be attributed to delayed diagnosis, advanced cancer at the time of detection, distant metastasis, and relatively limited access to advanced treatment technologies. Currently, TC diagnosis often requires ultrasound or pathology, however, due to the private nature of the location, ultrasound is not commonly employed as a routine screening method for early detection. Besides, cultural and religious factors in some Asian and African countries can delay diagnosis and treatment, as there may be reluctance to examine this area of the body. TC most frequently presents as a painless scrotal mass and testicular self-examination (TSE) has been shown to facilitate early detection, potentially improving patient outcomes and prognosis (44). Studies have demonstrated that promoting TSE as a low-cost screening tool can result in a gain of 46-50 life years for every TC death prevented (45). Nonetheless, the current uptake of TSE remains concerning (46). A study among men aged 18-35 years revealed limited awareness of TC and TSE, with only 17.6% of college students aware of how to perform TSE and the lowest utilization rates were observed in Southeast Asia (Singapore) (47). In addition to the lack of early screening and diagnosis, the unequal distribution of medical resources contributes to the high mortality rates in these regions. For instance, several Latin American countries, including Mexico, Brazil, and Cuba, continue to face shortages of medical resources and inadequate healthcare coverage (48), and India and Pakistan face similar healthcare challenges. Moreover, marijuana use, which is legal for religious purposes in some areas of India, may also be contributing to the rise in TC-related deaths (49).

### 4.3 Decomposition and frontier analysis of the SDI burden of TC in the AYA population

At the global level, the decomposition analysis reveals that population growth is the primary driver of the increased DALYs in the AYA population. In regions with high or high-middle SDI, epidemiological trends have resulted in a reduction in DALYs. In contrast, in the middle SDI region, the epidemiological trend is the main factor responsible for the higher DALYs, indicating that although countries in this region have relatively comprehensive incidence data for TC, the treatment infrastructure remains underdeveloped, leading to poorer patient outcomes. Furthermore, the middle SDI region contributes the largest share to global DALYs, highlighting the potential for improving TC diagnosis and treatment in these areas to substantially reduce global DALYs. Conversely, in regions with low SDI or middle-low SDI, population growth remains the primary factor driving the high DALYs observed in these areas.

The frontier analysis indicates that some middle and high-middle SDI countries, such as Mexico and Chile, are supposed to have more advanced medical technology and healthcare systems, but still exhibit high effective difference. Thankfully, Chile has enhanced cancer patient management in recent years with a comprehensive coverage program for 11 cancers containing TC, but there remains significant room for improvement (48). In contrast, certain lower SDI countries show low effective variances, which may be attributed to limited diagnostic and treatment technologies, as well as deficiencies in cancer registry data, causing suboptimal detection rates, yet demanding attention. For European and North American countries, such as the United States, Germany, and Canada, which face genetic and hereditary risks, DALYs have demonstrated a decreasing trend alongside a lower effective variance. This elucidates that these nations have well-established systems for the prevention, screening, and diagnosis of TC, positioning them as potential models for other countries burdened by high rates of testicular cancer.

### 4.4 The prevention, control, and management of TC in the AYA population

At the global level, the incidence and prevalence of TC in the AYA population continue to rise, pointing out that preventive measures remain insufficient. DALYs are also experiencing a gradual increase, reflecting ongoing improvements in diagnostic techniques and management of TC survivors. However, mortality rates continue to rise in certain countries and regions, underscoring the need for enhanced early screening, disease detection, and widespread adoption of standardized treatment protocols. For healthy individuals: i) Regular TSE should be conducted, particularly for individuals with a family history of TC. ii) Consumption of alcohol and tobacco should be reduced, and poor dietary and occupational habits should be addressed to minimize exposure to harmful substances. For TC survivors: i) patients should actively engage in follow-up care with medical institutions with no stigma related to this. ii) If issues related to fertility, sexual health, or emotional well-being arise, timely psychological counseling and professional treatment should be sought. For medical institutions: i) Continuously update diagnostic and treatment technologies to improve TC cure rates. ii) Establish standardized follow-up management programs for TC survivors, with a focus on fertility, psychological health, and cardiovascular issues. For governments: i) Enhance medical security systems and promote the equitable distribution of healthcare resources. ii) Given that the legalization of recreational marijuana may contribute to the rise in TC cases, relevant policies should be carefully developed and implemented. For the World Health Organization: i) Provide medical assistance and support to less developed regions and countries. ii) Improve relevant prevention, screening, and treatment systems in these regions.

## 5. Limitations

However, this study has several limitations. i) The GBD data do not provide specific pathological subtypes of testicular cancer. Different histological types of testicular cancer, such as non-seminomatous germ cell tumors, exhibit varying prognoses, which tend to be more aggressive to metastasize early (50). ii) The GBD database categorizes sex distinguishing only between males and females, so transgender populations may be underreported (51). Additionally, transgender individuals may face discrimination, negative healthcare experiences, and various forms of social pressure, which could lead to medical avoidance and subsequently lower detection rates. iii) In low-resource countries, limited coverage and inadequate quality of cancer registries may introduce bias in national-level data, affecting the accuracy and generalizability of the findings.

## 6. Conclusions

Overall, TC in the AYA population continues to impose a significant disease burden, influenced by genetic factors, environmental exposures, and poor lifestyle habits. Although the incidence and prevalence of TC have led to a decrease in DALYs thanks to advances in medical technology, this progress has also resulted in a growing number of survivors who face long-term issues such as infertility, mental health disorders, cardiovascular diseases, and other sequelae. Therefore, for the AYA population, it is essential to promote lifestyle modifications, such as reducing the consumption of marijuana, tobacco, and alcohol, as well as to encourage the widespread adoption of TSE techniques to facilitate early diagnosis. Furthermore, comprehensive follow-up care for TC survivors must remain a priority, requiring sustained efforts from governments to revise and implement social and medical policies to better address these challenges.

## 7. Ethics approval

Not applicable.

## 10. CRediT authorship contribution statement

Hongting Xie and Quan Sun: Conceptualization, Formal analysis, Writing-original draft. Peng Xue, Xuelei Chu, and Shijie Zhu: Conceptualization, Writing-original draft. Feiyu Xie and Lingze Xi: Formal analysis, Visualization, Data curation. Jiaming Hong, and Xingdong Lin: Formal analysis, Visualization. Jun Lyv: Methodology, Writing-review & editing. Nan Bu-Wang: Conceptualization, Formal analysis, Visualization, Writing-review & editing.

## 11. Availability of data and materials

All data are accessible through the GBD query tool (http://www.healthdata. org/gbd/2021).

## Data Availability

All data are accessible through the GBD query tool(http://www.healthdata.org/gbd/2021).

## 12. Acknowledgments

We highly appreciated the work of the Global Burden of Disease Study 2021 collaborators for providing the most comprehensive analysis of different diseases on a global scale.

## 13. Appendix A. Supplementary material

Supplementary data for this article can be found in the Appendix Supplementary Materials.

## 14. Abbreviations

AAPCs: Average annual percentage changes
ASR: Age-standardised rate
ASIR: Age-standardized incidence rate
ASPR: Age-standardised prevalence rate
ASDR: Age-standardised DALYs rate
AYA: Adolescents and young adults
CI: Confidence interval
DALYs: Disability-adjusted life-years
ED: Erectile dysfunction
GBD: Global Burden of Disease
SDI: Socio-demographic index
TC: Testicular cancer
TSE: testicular self-examination
UI: Uncertainty interval.

